# Family, community, institutional and policy factors on COVID-19 vaccine perceptions among urban poor adolescents in seven countries: qualitative cross-site analysis

**DOI:** 10.1101/2023.11.03.23298048

**Authors:** Astha Ramaiya, Kristin Mmari, Ana Luiza Borges, Cristiane Cabral, Eric Mafuta, Aimee Lulebo, Chunyan Yu, Anggriyani Wahyu Pinandari, Siswanto Agus Wilopo, Effie Chipeta, Kara Hunersen

## Abstract

**Purpose:** The number of studies examining family, community, institutional and policy factors on COVID-19 vaccine perceptions is limited, with most concentrating on high-income countries and using predominantly quantitative methods. To address this gap, the goal of this manuscript is to qualitatively explore these factors and how they shape adolescents’ perspectives on COVID-19 vaccines across seven countries.

**Methods:** Focus group discussions (FGDs) were conducted among urban poor adolescent populations (13 - 18 years) across seven countries: Ghent, Belgium; Sao Paulo, Brazil, Shanghai; China, Kinshasa, Democratic Republic of Congo (DRC); Semarang and Denpasar, Indonesia; Blantyre, Malawi and New Orleans, United States of America (USA). An inductive thematic analytical approach was used to understand the emerging themes across the different countries based on the study’s objectives.

**Results:** The study found that all influences were inter-connected and contributed towards vaccine perceptions among adolescents, which were largely positive except in the two African countries and to an extent in the USA. Family and community influences played a large role in vaccine perceptions, however, this differed by context. Our findings suggest adolescents’ perceptions about vaccines were more positive in countries with higher vaccination rates, i.e. China and Indonesia versus countries with lower vaccination rates i.e. Malawi and DRC. Vaccine mandates within schools, offices, and public places were also discussed with varying perceptions based on government trust.

**Conclusion:** Adolescents’ perceptions of the Covid-19 vaccine are based on a variety of elements, such as families, community, institutions, and policies. Prioritizing one or another path may not be sufficient to improve vaccine adherence during future pandemics, as we experienced with Covid-19. Strategies to make vaccine perceptions more positive among urban poor adolescents should address both family and community perceptions. However, policies and robust programs around immunization are still needed.

## Introduction

In March 2020, the World Health Organization declared COVID-19 a pandemic (1). After an expedited approval process facilitated by national and global health institutions, COVID-19 vaccines started rolling out among adults in December 2020 and among children and adolescents in 2022 (2). Currently, the World Health Organization (WHO) states that any individual should get any COVID-19 vaccine dose/s recommended by the country’s health authority including boosters when available (3). Along with other prevention methods such as mask-wearing and hand hygiene; vaccines are a safe and reliable method to build immunity against the virus, reduce the likelihood of severe illness and limit the transmission/acquisition of the virus (4). As of May 2023, 70% of the world has received at least one dose of a COVID-19 vaccine (5). However, this proportion significantly decreases in low-income countries (30%) due to issues including supply chain disruptions, lack of proper vaccine storage facilities, and vaccine hesitancy (5–7).

COVID-19 vaccinations during the adolescence period are critical for both personal and public health (8,9). Despite evidence of lower transmission and better individual health, there is a wide range in vaccine uptake across countries; ranging from 21% of adolescents (15 - 17 years) in Croatia to 92% of adolescents (12 - 17 years) in Argentina having received at least one dose of a vaccine (5). Like the phenomenon seen among adults, vaccination rates among adolescents are lower in low- and middle-income countries (LMIC) (32% in the Middle East), with geographical diversity in levels of vaccine hesitancy (10,11). These statistics must be considered with the stipulation that adolescents under age 18 need permission from a legal guardian for vaccination, thus limiting their self-efficacy in healthcare decision-making (8,10). There is ample data on adolescent perceptions of the COVID-19 vaccines around the world including fear of injections/side effects, the safety of the vaccine, misinformation, institutional/drug company distrust, perceived low illness vulnerability and preference for traditional remedies, all of which relate to the concept of vaccine hesitancy (11–14).

Vaccine hesitancy is defined as “the delay in acceptance or refusal of vaccination despite the availability of vaccination services” (15). Despite evidence that vaccines can contribute to achieving 14 out of 17 Sustainable Development Goal (SDGs) including ending poverty, reducing hunger, and reducing inequalities, vaccine hesitancy persists globally and was exacerbated during the COVID-19 pandemic (16). A systematic review outlined the determinants of vaccine hesitancy among adolescents including: 1) complacency or the perceived risk of contracting the disease; 2) convenience or access to vaccinations, physical availability, accessibility, affordability or quality of service; 3) confidence or trust in the effectiveness and safety of vaccine, trust in health care systems, motivations of policy makers; 4) contextual: historic, socio-cultural, environmental, health system/institutional, economic or political factors; 5) individual and group influence: personal perception of vaccine or social environment/peer; 6) vaccine specific issues that are directly related to the vaccine/vaccination (17). Conversely, a meta-analysis showed that interventions that include health education; vaccine mandates; provider education with performance feedback; class-based school vaccination strategy; multi-component provider interventions and multi-component interventions targeting providers and parents (including social marketing and health education) increase uptake of vaccinations among adolescents (18). Though much of the extant literature on adolescent acceptance of the COVID-19 vaccine focuses on individual and socio-demographic determinants (11,13,19–21), past reviews and literature on other types of vaccines have stressed the importance of understanding how all levels of the socio-ecological model drive vaccine perceptions (17,22,23).

Conformist social influence purports that people learn and adopt the behaviors of the majority, which can play a role in the acceptance of health innovations, which, in this case, is the COVID-19 vaccine (24). The increasing vaccination model shows that the interaction between an individual (perceived disease risk and vaccine confidence) and social factors (social norms, health worker recommendation, gender equity) motivates the individual to get a vaccination. However, practical issues (i.e., availability, affordability, ease of access, service quality, mandates, incentives and respect from health workers) moderate the relationship between motivation and vaccine uptake (25). During adolescence, social and institutional factors play a unique role in adopting healthy behaviors (25). In particular, what adolescents perceive is the behavior of others (descriptive norms) and the perception of other’s attitudes towards a behavior (injunctive norm) has been associated with the intent to take the Human Papillomavirus vaccine, Influenza vaccine, and COVID-19 vaccine among college students in the United States (26,27). Additionally, trust in the government plays an important role in both knowledge uptake and intent to follow recommendations and policies (28,29). However, the number of studies examining social and institutional factors on COVID-19 vaccine perceptions is limited, with most concentrating on high-income countries and using predominantly quantitative methods (12,14,22,26,27). There is a gap in the literature looking at the family, community, institutional and policy influences on vaccine perceptions among urban poor adolescents across different social contexts. To address this gap, the goal of this manuscript is to qualitatively explore these factors and how they shape adolescents’ perspectives on COVID-19 vaccines across seven countries.

## Methods

### Context

Eight urban adolescent populations across seven countries were included in the analysis: Ghent, Belgium; Sao Paulo, Brazil, Shanghai; China, Kinshasa, Democratic Republic of Congo (DRC); Semarang and Denpasar, Indonesia; Blantyre, Malawi and New Orleans, United States of America (USA). Data collection occurred between March 2021 to April 2022. As such, schools in all sites were reopened at the time of data collection following COVID-19-related closures, except in Belgium and Indonesia where schools were closed for the holidays.

Vaccines were permitted for all adolescents during the study period except in China and Malawi. Vaccine mandates were implemented in Brazil, Indonesia, Malawi, and the USA. In Brazil, schools mandated vaccinations; however, this was later revoked by the government (30). In Malawi, all frontline workers including health workers and journalists were required to get vaccinated (31). In Indonesia, all adults were mandated to be vaccinated and were fined or retracted from social assistance or government services if they were unvaccinated; vaccine certificates were also required to access public transportation (32). Lastly, in the USA, all federal workers, contractors, private sector workers with more than 100 employees, and public sector workers were required to get vaccinated (32). Currently, the proportion of the population vaccinated with at least one dose in each country ranges from 16% in DRC to 92% in China (5). Table 1 shows the contextual data.

**Table 1:** Country Contextual Data During Data Collection.

| Country | Data collection | School status | Were vaccines available for adolescents during data collection? | Vaccine mandates implemented [30–32] | Vaccines approved for use among adolescents (10 – 17 years)<sup>1</sup> | The proportion of country vaccinated with at least one dose (May 2023) [5] | Daily new confirmed COVID cases in the country during data collection [5] |
| --- | --- | --- | --- | --- | --- | --- | --- |
| Ghent, Belgium | March 2021 | Closed for holidays | Yes | No | Moderna<br>Pfizer/BioNTech | 79% | 2,376 - 4,840 |
| Sao Paulo, Brazil | September 2021 | Open | Yes | Yes | Pfizer/BioNTech<br>Sinovac (Coronavac) | 88% | 15,051 - 35,647 |
| Shanghai, China | June 2021 | Open | No | No | Sinopharm/ BIBP (Beijing)<br>Sinovac (CoronaVac)<br>Sinopharm/ WIBP(Wuhan)<br>Zhifei Longcom | 92% | 113 - 503 |
| Kinshasa, DRC | November 2021 | Open | Yes | No | Moderna<br>Pfizer/BioNTech<br>Janssen<br>Oxford AstraZeneca<br>Sinovac (CoronaVac) | 16% | 10 - 44 |
| Semarang and Denpasar, Indonesia | March - April 2022 | Closed for holidays | Yes | Yes | Sinovac (CoronaVac)<br>Oxford AstraZeneca<br>Sinopharm (Beijing)<br>Moderna<br>Pfizer/BioNTech<br>Novavax | 87% | 497 - 39,885 |

<sup>1</sup> Moderna and Pfizer/BioNTech are mRNA vaccines; Sinovac (CoronaVac), Sinopharm/BIBP (Beijing) Sinopharm/WIBP(Wuhan) use VeroCell to produce inactivated vaccine; Zhifei Longcom uses CHO cell to produce Recombinant Novel Coronavirus vaccine; Janssen and AstraZeneca are vector vaccines

| Country | Data collection | School status | Were vaccines available for adolescents during data collection? | Vaccine mandates implemented [30–32] | Vaccines approved for use among adolescents (10 – 17 years)<sup>1</sup> | The proportion of country vaccinated with at least one dose (May 2023) [5] | Daily new confirmed COVID cases in the country during data collection [5] |
| --- | --- | --- | --- | --- | --- | --- | --- |
|  |  |  |  |  | Janssen |  |  |
| Blantyre, Malawi | January – February 2022 | Open | No | Yes | Pfizer/BioNTech | 24% | 17 - 678 |
| New Orleans, USA | December 2021 | Open | Yes | Yes | Moderna<br>Pfizer/BioNTech | 81% | 97,313 - 310,812 |

### Data source

The Global Early Adolescent Study (GEAS) is a longitudinal study, initiated in 2012, on gender socialization and health among adolescents living in poor urban communities across nine countries (33). As part of the study, COVID-19 quantitative and qualitative modules were introduced among a sub-sample of adolescents in eight countries between 2020 and 2022. For this paper, we draw on qualitative data from the focus group discussions (FGDs) across seven countries that were part of the COVID-19 module. Quantitative data was not used because the variation in the proportion of adolescents being/willing to be vaccinated was too limited to make meaningful statistical associations with family, community, and institutional factors.

Except for Brazil and China, all sites used the same inclusion criteria for focus group recruitment, which included: 1) residing in an urban poor setting; 2) aged between 13 – 18 years; 3) and being a member of the GEAS cohort. In Brazil, since baseline data collection had only recently started, recruitment for the focus groups was not dependent on being a member of the GEAS. In China, older participants (aged 15-18) were not part of the GEAS cohort and were recruited from a nearby high school.

Data collection modality differed by country with Belgium, Indonesia, and USA conducting FGDs online, while Brazil, China, DRC, and Malawi conducted FGDs in-person. Topics discussed during the focus groups included COVID-19 knowledge and attitudes; sources of information; preventative practices; perceptions of the vaccines; the impact of COVID-19 on family, friends, school, economic and health; biggest concerns; coping mechanisms; and types of support needed. Questions about the vaccines were asked to participants only after vaccinations had rolled out in the country. Trained facilitators conducted the interviews. The FGDs were audio-recorded, transcribed, and subsequently translated into English for coding. Table 2 shows FGD stratification by country and data collection modality:

**Table 2:** Number and Modality of Focus Groups by Country.

| Country | Stratification and number of FGDs | Sample size in each group | Data collection modality |
| --- | --- | --- | --- |
| Ghent, Belgium | *Sex (n=2):*<br>1 Boy<br>1 Girl | Boys: 9<br>Girls: 13 | Online |
| Sao Paulo, Brazil | *Sex and mixed (n=7):*<br>1 Boy<br>1 Girl<br>5 Mixed | Boys: 5<br>Girls: 4<br>Mixed: 24 | In-person |
| Shanghai, China | *Sex and age (n=4):*<br>1 Older girls<br>1 Younger girls<br>1 Older boys<br>1 Younger boys | Older girls: 10<br>Younger girls: 10<br>Older boys: 10<br>Younger boys: 10 | In-person |
| Kinshasa, DRC | *Sex (n=4):*<br>2 Boys<br>2 Girls | Boys: 16<br>Girls: 14 | In-person |
| Semarang & Denpasar, Indonesia | *Sex and Socio-economic Status (SES) (n=8):*<br>1 Higher SES girls in Semarang<br>1 Lower SES girls in Semarang<br>1 Higher SES boys in Semarang<br>1 Lower SES boys in Semarang<br>1 Higher SES girls in Denpasar<br>1 Lower SES girls in Denpasar<br>1 Higher SES boys in Denpasar<br>1 Lower SES boys in Denpasar | Semarang:<br>Higher SES boys: 9<br>Higher SES girls: 10<br>Lower SES boys: 8<br>Lower SES girls: 8<br>Denpasar:<br>Higher SES boys: 6<br>Higher SES girls: 7<br>Lower SES boys: 10<br>Lower SES girls: 8 | Online |
| Blantyre, Malawi | *Sex and age (n=4):*<br>1 Older girls<br>1 Younger girls<br>1 Older boys<br>1 Younger boys | Older boys: 9<br>Older girls: 8<br>Younger boys: 7<br>Younger girls: 10 | In-person |
| New Orleans, USA | *Sex (n=3):*<br>2 boys<br>1 girl | Boys: 8<br>Girls: 4 | Online |

### Ethical Considerations

The study received ethical approval from the University of Ghent in Belgium; the University of São Paulo School of Public Health in Brazil; Shanghai Institute of Planned Parenthood Research in China; Kinshasa School of Public Health in DRC; Faculty of Medicine, Public Health, and Nursing, Universitas Gadjah Mada in Indonesia; College of Medicine Research Ethics Committee in Malawi; and Institute of Women and Ethnic Studies in USA. The study was approved for secondary data analysis in all countries except Kinshasa by the Johns Hopkins Bloomberg School of Public Health’s Institutional Review Board (IRB) (#8549). In Kinshasa, IRB approval was obtained for primary data collection from the Johns Hopkins Bloomberg School of Public Health (#7510). Written consent was obtained from adolescents who were 18 years old. Adolescents who were between 13 and 17 years old provided assent and parental consent.

### Analysis

An inductive thematic analytical approach was used to understand the emerging themes across the different countries based on the study’s objectives [44]. The transcripts were coded by two coders, who assured inter-rater reliability by coding the first four transcripts from round 1 together. Thereafter, the two coders independently coded the other 65 transcripts. A total of 32 transcripts had quotations on vaccination. Once the coding was completed, within- and cross-country matrices were created for each code based on FGD stratification. All analyses were conducted on Atlas.ti 9.1 [45].

## Results

We organized factors by socio-ecological domain (family, community, institutional, and policy) to reflect how these various contextual levels all impact adolescents’ perceptions on vaccine trust and hesitancy across the different countries.

### Family factors

Attitudes of family members, as well as their own personal experiences with the vaccine, played an important role in vaccine perceptions. Interestingly, the way in which these factors influenced adolescents largely depended on the setting. For example, in Indonesia, adolescents were more likely to trust the COVID-19 vaccine as a result of their parent’s positive experiences with the vaccine.

> “In my opinion, vaccines are mandatory and must be done because of the experience of parents who got it, [they’ve] already been vaccinated twice, the symptoms are lighter and for example, if we get vaccinated too, we can go out of town like that. You have to get vaccinated, right? If it’s from me.” (Boy, higher income, Denpasar, Indonesia)

In contrast, in the DRC and Malawi, it was discussed in context of vaccine hesitancy. Girls in the DRC talked to a greater extent about adverse events occurring to their family members after taking the vaccine which influenced their perceptions:

> “Because our grandmother is in Europe, she wanted to come here, she had already sent the goods, she received the first wave, first dose, it had made her fever and the second dose when she had gone to take when she went back to wash herself, she had become black all the body until the day when the person itself will die. (How do you know it’s related to the vaccine?) because she was fine when she took it, (but now will you take it?) I won’t take it.” (Girl, DRC)

In Brazil, adolescents talked about mothers’ requiring the adolescents to get the vaccine irrespective of what was recommended. However, adolescents also criticized the distrust of some family members:

> “No, my mother was very insistent that I should go, she came and said “yes, you are going to be vaccinated”, it was me, my mother, everyone from home, fine.” (Mixed group, Brazil)

> “Then she[grandmother] thinks she is not going to get the vaccine, and I say “where is your sister to take care of you?” Because her sister keeps putting in her head that this vaccine is from the devil, that it’s this, that it’s that. Then I said “well, now you call your sister to take care of you, because you don’t listen to us”.” (Mixed group, Brazil)

The USA was unique in that adolescents had mixed attitudes about the vaccine and its effectiveness, which was influenced by family members’ experiences:

> “Well like, for instance, my mama is scared of getting the vaccine because she thinks it’s gonna make us even sicker. So, I don’t think that getting the vaccine is a bad thing because my grandma got it and she’s an old person and COVID is like really dangerous to the old people. So I don’t really think it’s that bad.” (Girl, USA)

> “I don’t know if I trust it or don’t trust it because I just, I really don’t even know what to think about it. Like, some of my family members. They got it. But my aunt, she had COVID first. And then she waited like, three months or six months later to get the shot. She says she feels regular. So, and I wouldn’t know.” (Girl, USA)

### Community factors

Community influences, such as community trust and social cohesion, also shaped vaccine-accepting perceptions. Adolescents in China, in fact, reported strong community trust and influence in getting the vaccine.

> “We are all responding to the call of the community. People, including teenagers like us, want to get the vaccine.” (Girl, younger, China)

In DRC and Malawi, descriptive norms played a role in being vaccine hesitant. Adolescents described that they didn’t see other community members getting the vaccine; this was often coupled with misinformation that was acquired from the community:

> “If I’m told to take the vaccine, I won’t take it because since the vaccine has been around, I haven’t seen anyone get vaccinated, and I wouldn’t like to be the first one to be vaccinated, in our community, in our neighborhood, I haven’t seen anyone.” (Girl, DRC)

> “What people said is that when you get the vaccine and you tested negative, you will get sick and then you die.” (Girl, Older, Malawi)

However, there was some discussion in DRC about trusting doctors and observing reduced mortality within the community; as well as the church influencing positive vaccine perceptions:

> “We used to wear the muffler because there was no hope but now with the vaccine there is hope and our doctors can treat this disease. The rate of the disease has gone down, out of 100% you can get even 45%.” (Boy, DRC)

> “I will accept to take the vaccine because I have already seen many people who have been vaccinated, especially in our church, so I will accept. The vaccine will protect me and those around me.” (Girl, DRC)

In Malawi, girls and boys noted that pastors and the church were advising against the vaccine:

> “Lately people have been complaining about the vaccine. Some pastors have been preaching against it saying it is satanic although some claimed it is good. So we don’t know really whether it is good or bad.” (Boy, older, Malawi)

### Institutional factors

Institutional trust played a large role in vaccine perceptions. In Indonesia and China, adolescents trusted the government because they believed it protected themselves and the community. In China, adolescents compared the COVID-19 mitigation measures to the rest of the world:

> “In my opinion, the current epidemic prevention and control in China is relatively perfect, and the Chinese government’s current hope is to vaccinate everyone, vaccinate as much as possible, so that the effect of total immunization can be achieved. Well, plus, the Chinese overall economic level is at the forefront compared with the international level, and this situation is relatively optimistic” (Boy, older, China)

> “the vaccine is really very good at keeping us protected from this covid 19. It’s not protected to prevent the symptoms. So, if the government imposes or obliges us for the COVID-19 vaccine, it really must be done because the Covid-19 vaccine is as important as that.” (Girl, lower income, Denpasar, Indonesia)

The fact that the vaccine was rapidly developed acted in both directions, both to foster belief in the vaccine and to distrust it. In Belgium, most adolescent girls discussed trusting the vaccine because it was rigorously tested by scientists. However, some girls expressed frustration they were not able to access the vaccines:

> “Because all the world was working on a vaccine against corona, everyone was only making the vaccine so it had to be something that would protect us anyway if it did not work it would not have passed those tests.” (Girl, Belgium)

> “with us in Bredene the vaccination center is ready but Bredene and De Haan have to work together, but the problem is for us, no one in the whole of Bredene has had an email to say when they can go or really any info at all…The thing is, it is ready and there are signs everywhere where you need to go, but you actually don’t know anything about it” (Girl, Belgium)

In Brazil and Belgium, adolescents voiced skepticism of the “effectiveness of a product developed so quickly in the history of a scientific research”. Whereas in Brazil, it was discussed within the context of some close family members to get vaccinated; in Belgium it was among adolescents themselves:

> “Ah, my family, some were not vaccinated. My uncle says that there was no time to create a vaccine, and because the president said he was not going to take it, and that’s it, he didńt took it.” (Girl, Brazil)

> “Madam, I don’t think it is really reliable because they found the vaccine already n one year so I think it is a bit of a fast period and there is also a discussion that there may be other substances in the vaccine so that the world population cannot increase or that women cannot get pregnant so that it does not rise or something” (Girl, Belgium)

In Malawi and DRC, vaccine hesitancy was discussed in the context of government and healthcare worker mistrust:

> “If the vaccine can be made open to everyone I cannot go and get it because of the rumors that spread saying that it is just meant for people in authority to benefit financially.” (Boy, older, Malawi)

> “He himself works in VIJANA [health center], he himself sends us to take but he himself has not taken yet but in other hospitals, they require other people to take only the shot but they themselves do not take” (Girl, DRC)

Adolescents in Malawi and DRC also talked about how media (TV channels, radio, and social media) played a role in vaccine mistrust and misinformation. This was discussed to a greater extent by girls in comparison to boys and extensively discussed in DRC.

> “… this vaccine is not good, its effects are very dangerous, some say that it deforms, it gives high fever, and it kills people. Facilitator: where did you get all this information? Participant: In some TV channels, on the radio and even on social networks is what scares me.” (Girl, DRC)

### Policy factors

Vaccine mandates and associated perceptions were discussed in Brazil, China, Indonesia, Malawi, and the USA. These perceptions were usually linked to government trust and whether adolescents believed that vaccine mandates were protective.

In Brazil and Indonesia, adolescents talked about requiring vaccinations to enter school and attending in person classes:

> “The students didn’t take the vaccine in the beginning, they did it again, it was mandatory, now the teachers have already taken it… That would be the right thing, everyone should get vaccinated, at least the students who are the majority instead of the teachers.” (Girls, Brazil)

> “Is there anyone of you who hasn’t been vaccinated? no, we already have been…because it’s mandatory from school, I guess.” (Girl, higher income, Denpasar, Indonesia)

In China and Indonesia, adolescents discussed vaccines’ being required to go to the office (for family members), accessing public spaces, and traveling. Adolescents in these two countries were also more in favor of the mandate because it allowed them to move around:

> “My family all got the vaccine. There are some KPI [key performance indicators] in their office, and then they are forced to do so. That is to say, every time their company will pull a performance table of their department, and then they will ask those who have been vaccinated to tick it, and every day they would call those who have not been vaccinated.” (Girl, older, China)

> “Because it’s easy to go anywhere, because yesterday you had to be vaccinated if you wanted to go anywhere. You have to have a vaccine card, a vaccine certificate or something. So make it easier to do anything.” (Girl, lower income, Denpasar, Indonesia)

In Malawi, vaccine mandates were associated with job loss. Most adolescents mentioned that because their parents didn’t get the vaccine, they were fired from their jobs and thus lost household income:

> “Covid-19 affected businesses. People stopped going out to do business and because of Covid-19 people were being forced to get vaccines. As such it affected our fathers who refused to get the vaccine because they were sacked and had nothing to do for survival.” (Boy, older, Malawi)

In Indonesia, most adolescents talked positively about how vaccine mandates helped to increase the uptake of vaccines in the community. However, some adolescents in Indonesia infringed on their rights:

> “Because as I said earlier, the vaccine is really very good at keeping us protected from this covid 19. It’s not protected to prevent the symptoms. So, if the government imposes or obliges us for the COVID-19 vaccine, it really must be done because the Covid-19 vaccine is as important as that, Miss.” (Girl, lower income, Denpasar, Indonesia)

> “It shouldn’t be too pushy, because it’s your own will. So human rights are enforced, they can’t be forced like that, especially if those who come to their house are forced to do that.” (Girl, lower income, Semarang, Indonesia)

In USA, adolescents were skeptical about the vaccine mandates because they didn’t think it was effective and because they thought that these were infringing on their human rights. In some instances, family perceptions of the mandates influenced adolescent’s perceptions:

> “I feel like it’s going to affect the pandemic because you know, like the government and everybody I feel like they just gon’ try to pressure you into getting the vaccine you know, like, try to enforce it into other places you know, like you can’t go here if you don’t have the vaccine and stuff like that, like I just feel like they’re gonna try to force you upon getting a vaccine.” (Girl, USA)

> “I don’t know how it will effect my household because we are not vaccinated because my grandma believes crazy things like the government Is trying to control us” (Boy, USA)

## Discussion

The purpose of this manuscript was to qualitatively look at the family, community, institutional, and policy influences on vaccine perceptions among urban poor adolescents across seven countries. Family and/or community influences played a large role in vaccine perceptions, however, this differed by context. Our findings suggest adolescents’ perceptions about vaccines were more positive in countries with higher vaccination rates, i.e., China and Indonesia versus countries with lower vaccination rates i.e., Malawi and DRC (34). Vaccine mandates within schools, offices, and public places were also discussed with varying perceptions based on government trust. With the pandemic transitioning to an endemic disease, COVID-19 vaccines will likely be integrated into routine immunizations to reduce hospitalization and mortality [46]. This research can contribute towards evidence on how family, community, institutional and policy influences are associated with vaccine perceptions yet interdependent within urban poor settings. Furthermore, this evidence can play a role in tailoring messages to reduce vaccine hesitancy for future vaccination programs including pandemics.

Perceptions of vaccines varied by geographical setting. This was particularly highlighted in the Sub-Saharan countries. A history of structural and systematic inequalities with a lack of culturally informed research looking at factors of vaccine hesitancy (35) contributes to why DRC and Malawi had predominantly negative perceptions about COVID-19 vaccines. These perceptions were usually coupled with misinformation and government mistrust. Research in the continent has shown that public distrust of the COVID-19 vaccine was because of concerns with vaccine safety and side effects; lack of trust in pharmaceutical industries; and misinformation/conflicting information from the media (36). Additionally, the delay in COVID-19 vaccine implementation in the continent coupled with vaccine trial abuse in the past has led to further vaccine hesitancy in the continent (35). In order to address future vaccine hesitancy within the continent, the literature recommends that: 1) Africa CDC and WHO Africa Regional Office coordinate, engage and mobilize communities to reduce misconceptions and determine future vaccine roll-out strategies; 2) WHO should share the lessons learned from the social mobilizations and communication campaigns for future vaccination campaigns; 3) Every country within the continent should create an office under the umbrella of WHO and Africa CDC to address fake news and misinformation; 4) Resources should be provided by international funders to support both the logistics and human resources to implement health promotion programs (35). The rollout of the Ebola vaccine in West Africa was deemed a success because communities were engaged in the trial from the beginning based on the principles of the four R’s: reciprocity, relatability, relationships, and respect. Both the local community liaison teams and the social science teams engaged and incorporated community member suggestions to improve the trial as well as foster trust in an egalitarian manner (37).

Family and community factors played an important role in vaccine perceptions across all the sites. We found the vaccination status of the family and/or community; vaccine experiences of the family and/or community; and re-infection of family/community members after getting the vaccine were all factors affecting vaccine perceptions among adolescents. This is important because adolescents are not autonomous to take their vaccine and they need their parents’ support and motivation to do so (8). Therefore, strategies to reach adolescents are not sufficient if they do not engage parents and communities. A scoping review showed that people would be more hesitant if they knew someone with a serious vaccine reaction, or not knowing someone close affected by COVID-19 (22). There is a need to address family and community factors in vaccine campaigns through social mobilization to promote vaccine demand (38). This can be through equipping trusted people from the local context to foster community ownership of the campaign; and investing more in social mobilization with linkages to service delivery to reduce frustration (38). Further research is also required to understand if adolescents’ perceptions on vaccines would motivate family members to get a vaccine.

Government trust, media and vaccine mandates were all factors affecting vaccine perceptions among adolescents across most countries. For example, in Indonesia, travel restrictions were enforced very hard and usually coupled with the government media publishing epidemic numbers, which is why perceptions of vaccines and mandates were largely positive. Past studies have shown that trusting the government is associated with adhering to policies proposed by the government (39–41). A study across 177 countries showed that a higher level of government and interpersonal trust together with lower government corruption was associated with higher vaccine coverage and lower infections (42). However, exposure to social media has shown to increase vaccine hesitancy (22). In order to address future pandemics/crises, adults, including governments need to address or maintain public trust by providing accurate and timely information, combating disinformation and misinformation online, getting vaccinated to reinforce trust and including social mobilization as a mechanism to improve health behaviors (42,43).

This study had limitations that need to be addressed. We used a purposive sample from urban poor settings and part of an existing cohort which could introduce selection bias (i.e.: those who volunteered to participate and those who did not). Stratification as well as focus on COVID-19 (i.e. epidemiological figures, media and public exposure) differed by site which affected the extent to which we could make comparisons across sites. The translation, transcription, and back translation process was conducted by the local researchers in each country. However, every facilitator and transcriber were trained before data collection. The data collection modalities and discussion around vaccines differed across sites which could affect the type of information obtained and the extent to which the results are generalizable. Despite these limitations, this study is strengthened by the fact that it explored several factors related to perceptions of COVID-19 vaccine hesitancy and acceptance across a wide variety of different cultural settings.

## Conclusion

Adolescents’ perceptions of the Covid-19 vaccine are based on a variety of elements, such as families, community, institution, and policies, which are all interconnected. Prioritizing one or another path may not be sufficient to improve vaccine adherence during future pandemics, as we experienced with Covid-19. Strategies to make vaccine perceptions more positive among urban poor adolescents should address both family and community perceptions. However, policies and robust programs around immunization are still needed. Mandates are still contradictory since they may improve vaccine adherence but raise concerns about free choices and rights.

It is important to emphasize that the distrust or hesitation surrounding the vaccine against COVID-19 seems to be just another element that makes up a current pandemic scientific denialism. Robust public policies are needed to face these denialist movements that produce disinformation on a large scale with the dissemination of fake news, as well as to restore the trust once deposited in public health institutions.

## Data Availability

The data is available as a Supporting File.

## Conflict of Interest

The authors declare no conflict of interest.

## Acknowledgements

The authors would like to acknowledge all the program staff in all of our countries: University of Ghent; University of São Paulo School of Public Health; Shanghai Institute of Planned Parenthood Research; Kinshasa School of Public Health; Universitas Gadjah Mada; Kamuzu University of Health Sciences (formerly College of Medicine) and Institute of Women and Ethnic Studies who worked under challenging circumstances to collect this data during a pandemic.

Additionally, we would like to acknowledge the GEAS Hopkins Coordinating Center who analyzed the cross-country qualitative data. Lastly, we would like to give our greatest appreciation to our participants who provided us with such rich and intriguing data.

## Funding

This work was supported by a gift agreement between AstraZeneca Young Health Programme and Johns Hopkins University.

## Footnotes

1 Moderna and Pfizer/BioNTech are mRNA vaccines; Sinovac (CoronaVac), Sinopharm/BIBP (Beijing) Sinopharm/WIBP(Wuhan) use VeroCell to produce inactivated vaccine; Zhifei Longcom uses CHO cell to produce Recombinant Novel Coronavirus vaccine; Janssen and AstraZeneca are vector vaccines

## Notes

### Competing Interest Statement

The authors have declared no competing interest.

### Funding Statement

This work was supported by a gift agreement between AstraZeneca Young Health Programme and Johns Hopkins University. The funder did not play any role in the submission.

### Author Declarations

The study received ethical approval from the University of Ghent in Belgium the University of São Paulo School of Public Health in Brazil Shanghai Institute of Planned Parenthood Research in China Kinshasa School of Public Health in DRC Faculty of Medicine, Public Health, and Nursing, Universitas Gadjah Mada in Indonesia College of Medicine Research Ethics Committee in Malawi and Institute of Women and Ethnic Studies in USA. The study was approved for secondary data analysis in all countries except Kinshasa by the Johns Hopkins Bloomberg School of Public Health’s Institutional Review Board (IRB) (#8549). In Kinshasa, IRB approval was obtained for primary data collection from the Johns Hopkins Bloomberg School of Public Health (#7510).

